# An Analysis of household catastrophic health expenditure and food insecurity in Ghana

**DOI:** 10.1101/2025.03.31.25324988

**Authors:** Jones Baah-Abekah, Duah Kwaku Ibrahim, Thomas A. Atugba, Seke Theodore, Shiddo Adamu Seidu, Donald Casley-Hayford, Larbie Deborah, Boaz Ahulu

**Author notes:** Corresponding author’s.

## Abstract

**Introduction:** Catastrophic health expenditure (CHE) occurs when healthcare expenses exceed 10% or 25% of household income, or health expenses exceed 40% of non-food household expenditure. The incidence of catastrophic health expenditure in Sub-Saharan Africa (SSA) is between 12.9% and 20.4%. The relationship between CHE and food insecurity has been identified, but the literature on SSA and Ghana is sparse. Against this backdrop, concerted efforts are needed to identify the driving factors behind CHE and food insecurity. The high rate of OOP payments poses substantial risks to financial stability for many families, particularly those from lower socioeconomic backgrounds. Addressing these issues will ensure all Ghanaians can access quality health services without financial strain. This study analyzed the health spending of households and their relationship with food insecurity in the Eastern region of Ghana.

**Objectives:** We examined the prevalence of household CHE and the relationship between CHE and food insecurity in the Eastern region of Ghana.

**Methods:** The study objectives were to estimate the prevalence of CHE in the Eastern region of Ghana and its association with food insecurity. This study analyzed secondary data from the Ghana Annual Household Income and Expenditure Survey (AHIES), conducted quarterly by the Ghana Statistical Service (GSS) between 2021 and 2023, using a subset of data for the Eastern Region of Ghana. We retrieved data comprising 5990 households and 52 variables from the GSS website and exported it to R for statistical analysis. Relevant variables important to the study’s objectives were identified and aggregated at the household level. To determine our predictors, we utilised forward stepwise logistic regression. A variable was considered significant to the model if its inclusion decreased the model’s Akaike Information Criterion (AIC). Any variables that caused an increase of more than 2 points in the AIC were omitted from the model. After determining the final model, we applied it using the 25% and 40% household income thresholds.

**Results:** Among the households that had expended on health in the 2 weeks before data collection, 985 (16.4%) households experienced CHE at 10% relative to total household income, 929 (15.5%) at 25%, and 903 (15.1%) at 40%. The prevalence of households experiencing food insecurity among those who had utilised health services was 2,121 (61.9%). CHE was significantly associated with household food insecurity at all levels. Households experiencing CHE at 10% of household income were significantly more likely to experience food insecurity as well, OR (95% CI; p) = 1.76 (1.51, 2.07; p<0.001). Households in rural areas were also more likely, 1.47 (95% CI 1.25, 1.72; p<0.001) at 10%, 1.53 (95% CI 1.30, 1.80; p<0.001) at 25%, and 1.60 (95% CI 1.35, 1.89; p<0.001) at 40% of household income, to experience CHE, all statistically significant. Households with a member aged over 60 years or under 18 years old had higher odds of CHE, with an OR of [1.45 (95% CI 1.22, 1.71; p<0.001)] and [1.32 (95% CI 1.09, 1.61; p=0.004)] at a 10% threshold income, respectively.

**Conclusion:** There is a relatively high prevalence of Catastrophic Health Expenditure, which is significantly associated with food insecurity at the 10%, 25%, and 40% thresholds of household income in the Eastern region of Ghana. The educational level of the household head, the presence of a minor or older household member, and urbanicity are predictors of CHE. The national health insurance scheme is insufficient in providing financial risk protection, and the government must revisit its implementation.

## Introduction

According to the World Health Organization (WHO), global efforts towards universal health coverage (UHC) have stagnated since 2019 and are not likely to be achieved by 2030 as expected. (1). The WHO notes that while the COVID-19 pandemic played a role, evidence suggests targets would have been missed regardless. As a result, more than half the world’s population lacks health coverage, and a further 2 billion people risk Catastrophic Health Expenditure (CHE), Impoverishing Health Expenditure (IHE), or both. (1). CHE occurs when healthcare expenses exceed 10% or 25% of household income (2,3), or health expenses exceed 40% of non-food household expenditure (4,5). The incidence of catastrophic health expenditure in Sub-Saharan Africa (SSA) is between 12.9% and 20.4% (6). The estimate is comparable to countries on other continents. For instance, in Bangladesh, 14.2% has been reported (7). Factors such as the lack of health insurance, lower educational attainments, suffering from chronic disease, elderly household members, Out-Of-Pocket payments (OOP), and rural residence are the main drivers of CHE (7–11). Despite health insurance coverage providing some protection against CHE, there is ample contrary evidence that it is insufficient for most families (12– 14)Even though a few studies have found evidence that health insurance coverage prevents CHE (15,16). High OOP has been identified as the main driver of CHE, and many countries have attempted to minimise OOP healthcare expenditure. In the face of CHE, households resort to denying themselves essential livelihoods such as food, education, and further healthcare. Households compensate by taking credit or disposing of assets (including income-generating assets) and using household savings (16–19).

The government of Ghana’s expenditure on health has consistently been below the Sub-Saharan African average despite its commitment to the Abuja declaration for the government to spend 15% of the national budget on health. The government of Ghana currently spends about 4.2% of its Total Gross Domestic Product on healthcare, which is lower than other sub-Saharan African countries (20). This trend raises significant concerns regarding the sustainability of health financing and its implications for service delivery, especially for vulnerable populations who disproportionately bear the burden of healthcare costs. Thus, Ghana’s commitment to Universal Health Coverage by 2030 remains in jeopardy due to the disparities in healthcare access and the high OOP expenses that many households face, including payments for services stated to be explicitly free (21).

Ghana introduced the National Health Insurance Scheme (NHIS) through the Parliamentary Act 650, and it evolved in 2012 from a district mutual health insurance to a national health insurance program. Despite a decade of NHIS in Ghana, enrollment in the scheme remains at only 67% coverage (22); OOP remains very high in Ghana, accounting for 27.2% of all health expenditures (23). A 2019 study found the average OOP during health for individuals to be GHS 27.04 (USD 5.55) (24). Most households cited the cost of the insurance premium as the reason for not enrolling in the NHIS; other factors such as income, socio-economic status (SES), formal employment, and educational status have been identified to determine household enrollment in health insurance significantly. Despite this, some studies have found that NHIS membership protects households and individuals from CHE (21,25,26). However, contrary evidence shows that its protection is minimal (27,28). A systematic literature review estimated that 30% of households in Ghana experience CHE (29), but a study in 3 communities in Ghana found CHE to occur in 11.4% of healthcare spending, at 20% of household income (28) The survey by Kusi et al. (2015) is similar to studies in neighboring countries such as Burkina Faso (30). A disease-specific estimate put CHE at 45% among patients seeking surgical care (27) and 19% for women accessing skilled delivery services (31), suggesting there are nuances to the incidence of CHE in Ghana.

Food insecurity refers to a condition where individuals or groups do not have consistent access to enough safe and nutritious food for healthy growth and development. This situation often arises from a combination of factors, including the unavailability of food and insufficient resources to buy it.(32). While literature exists on food insecurity in Ghana (33–40), none to the best of our knowledge), discusses its effect on healthcare expenditure, except Daniel (2024), who explored the role of food security status on the NHIS (38). The relationship between CHE and food insecurity has been identified (17,41,42)However, the literature on SSA and, specifically, Ghana is sparse. Against this backdrop, concerted efforts are needed to identify the driving factors behind CHE and food insecurity. Knowledge of household health expenditure patterns is crucial for informing policies to enhance access to healthcare, achieve UHC in Ghana, and prevent food insecurity. The high rate of OOP payments poses substantial risks to financial stability for many families, particularly those from a lower socioeconomic background. (7,43). Addressing these issues will ensure all Ghanaians can access quality health services without financial strain. This paper analyses the health spending of households in the Eastern region of Ghana and its relationship with food insecurity.

## Methodology

### Study setting

The Eastern Region is Ghana’s sixth-largest region, encompassing approximately 8.1% of the country’s total land area. With an estimated population of 2,633,154, it ranks as Ghana’s third-most populous region. Agriculture is the primary economic activity in the region, followed by industry. The region also has numerous traditional authorities, predominantly of Akan descent. There are 22 districts within the Eastern Region, most of which are rural (44).

A distinctive feature of this region is its strategic position between the Greater Accra Region and the Ashanti Region, the two most economically vibrant areas in Ghana. Analysing the Eastern Region provides valuable insights into the dynamic interplay of urban and rural households in Ghana.

### Data extraction

This study analysed secondary data from the Ghana Annual Household Income and Expenditure Survey (AHIES), conducted quarterly by the Ghana Statistical Service (GSS) between 2021 and 2023 (44)Based on the population and housing census data from 2021, the GSS sampled 10,800 households from 304 rural and 296 urban areas (enumeration areas). The GSS further sampled 18 households from each enumeration area to ensure regionally representative expenditure data for the Gross Domestic Product. This paper analyses the subset of data from participants in the Eastern Region of Ghana. The data was retrieved from GSS’s website and exported to R for statistical analysis. Relevant variables to the study were identified and aggregated at the household level for analysis. We created a variable to indicate whether there were household members less than 18 years, another to indicate whether there were household members more than 60 years, and lastly, the percentage of household members with active insurance coverage.

We initially calculated household income by estimating a monthly wage for each household member based on all sources of earnings and employment if a person had multiple income sources. In cases where an individual’s earnings were not monthly, e.g., yearly income, the monthly earnings were computed by dividing the annual earnings by 12 to obtain a standardised monthly income. We summed each household’s income and health expenditures and created binary variables to determine if spending exceeded income by 10%, 25%, or 40%, as defined in similar studies [2–5].

Data on household food security were already aggregated at the household level. A household was classified as experiencing food insecurity if any family member indicated they were worried about not having enough food, did not eat at all, or only consumed limited food due to a lack of money. Eight such variables were identified based on this classification. For example, a household was deemed to have experienced food insecurity if members of the household positively responded in the affirmative to any of the questions.

### Statistical analysis

A dataset comprising 5990 households and 52 variables was analysed. Summary statistics for each variable, including counts and percentages for categorical variables and means (with standard deviations) or medians (accompanied by the 25th and 75th percentiles) for continuous variables, as appropriate, were presented. Forward stepwise logistic regression was utilised to determine the predictors for the study. A variable was considered significant to the model if its inclusion decreased the model’s Akaike Information Criterion (AIC). Variables that caused an increase of more than 2 points in the AIC were omitted from the model. After determining the final model, the best-fitting model based on the AIC was applied to the 25% and 40% thresholds.

## Results

### Household characteristics

Most households lived in a rural area, 3,336 (56%), with a median (Q1, Q3) household population of 4 (3, 6). There were 2,252 (38%) female-headed households, compared to 3,265 (55%) male-headed households. The rest were single-member households. Most households, 4,587 (77%), had a minor, defined as a household member less than 18 years, and the median (Q1, Q3) median household age was 21 (14, 33). Older household members averaged a median (Q1, Q3) age of 49 (38, 62). There were 1,652 (28%) households that had a person more than 60 years old. Most household heads did not have a high school education, 3,170 (53%), with 1,997 (33%) having a high school education and only 802 (13%) households having a post-high school education (University, professional training, etc.) (Table 1).

**Table 1:** Household characteristics.

| Characteristic | N = 5,990<sup>1</sup> |
| --- | --- |
| Urban/Rural |  |
| Urban | 2,654 (44%) |
| Rural | 3,336 (56%) |
| Family size | 4 (3, 6) |
| Head of household |  |
| Female-headed | 2,252 (38%) |
| Male headed | 3,265 (55%) |
| Single | 473 (7.9%) |
| Child < 18 years | 4,587 (77%) |
| Median household ages | 21 (14, 33) |
| Ages of the oldest household members | 49 (38, 62) |
| Household member >60 years | 1,652 (28%) |
| Highest educational level |  |
| No high school education | 3,170 (53%) |
| High school education | 1,997 (33%) |
| Post-high school education | 802 (13%) |
| Unknown | 21 |

<sup>1</sup>n (%); Median (Q1, Q3)

### Household Health Protection and Health Services Utilization

The mean (SD) National Health Insurance coverage per household was 58 (39.3), with 2,060 (34%) households having a member with an additional free care or a subsidised health care plan from their employment. The mean (SD) of days during which a household member had been unwell was 5 (7.5), with a mean (SD) of 3 (6.0) being the number of days household members had taken off work. For households that assessed a medical service, 58 (1.0%) sought an obstetric service (antenatal, delivery, post-natal), 82 (87%) were for an acute service, and 12 (13%) were for check-up/investigative services. 657 (11%) households sought alternative health services like herbal, acupuncture, etc. The mean (SD) of total household income was GHS 720 (2,959.4) (Table 2).

**Table 2:**
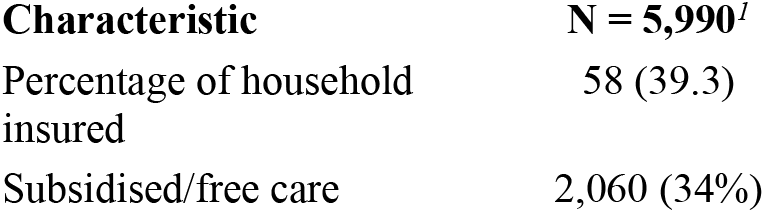
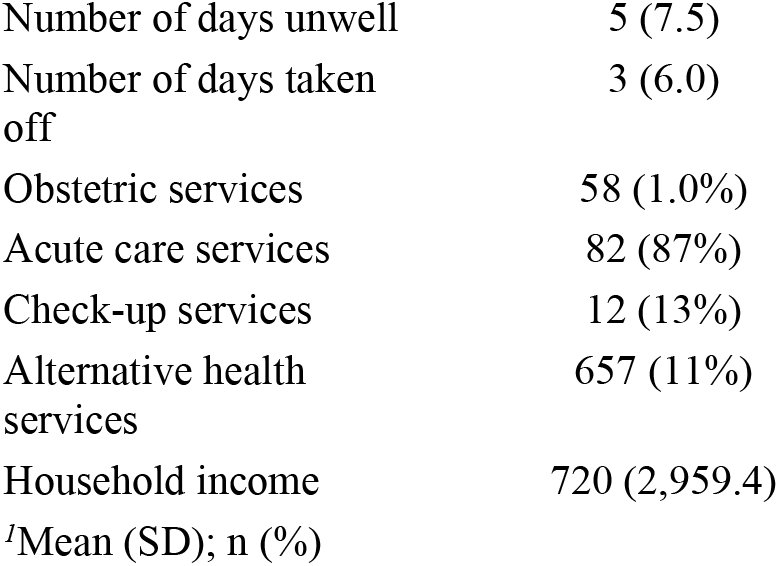
Health protection and health utilisation 2 weeks before data collection.

### Household income and health expenditure

In the last 2 weeks before data collection, the mean (SD) health expenditure by households for registration/card/folder was 2.55 (6.50), 6.37 (21.34) for consultation services, 14.78 (85.70) for diagnostic services (laboratory, x-ray, etc.), and 61.65 (149.57) on drugs. All (registration, consultation, services, and medicines) put together, the mean (SD) household treatment cost was 84.92 (201.39). The mean (SD) household expenditure on other medical services was 5.41 (49.66), and 10.93 (26.77) was spent on traveling for healthcare services on average. Households paid a mean (SD) of 11.76 (61.01) on admission and 51.56 (117.91) on medical supplies. Overall, the mean (SD) health expenditure per household was 158.84 (316.44) (Table 3).

**Table 3:** Household income and expenditure in the last 2 weeks before data collection.

| Characteristic | N = 1,747<sup>1</sup> |
| --- | --- |
| Registration | 2.55 (6.50) |
| Consultation | 6.37 (21.34) |
| Diagnoses | 14.78 (85.70) |
| Drugs | 61.65 (149.57) |
| Treatment cost | 84.92 (201.39) |
| Services | 5.41 (49.66) |
| Travel | 10.93 (26.77) |
| Admission cost | 11.76 (61.01) |
| Medical supplies | 51.56 (117.91) |
| Average health expenditure | 158.84 (316.44) |
| <sup>1</sup>Mean (SD) |  |

### Catastrophic health expenditure and food insecurity

Among the households that had expended on health in the 2 weeks before data collection, 985 (16.4%) households experienced CHE at 10% relative to total household income, 929 (15.5%) at 25%, and 903 (15.1%) at 40%. The prevalence of households experiencing food insecurity among those who had utilised health services was 2,121 (61.9%) (Table 4).

**Table 4:** Catastrophic health expenditure at 10%, 25%, and 40% of household income.

| Characteristic | N = 5,990<sup>1</sup> |
| --- | --- |
| CHE at 10% | 985 (16.4%) |
| CHE at 25% | 929 (15.5%) |
| CHE at 40% | 903 (15.1%) |
| Food insecurity |  |
| No risk | 1,303 (38.1%) |
| Food insecurity | 2,121 (61.9%) |
| <sup>l</sup>n (%) |  |

### Association between CHE and food insecurity

CHE was significantly associated with household food insecurity at all levels. The odds of experiencing CHE at 10% of household income were significantly more likely to experience food insecurity, with an odds ratio of OR (95% CI; p) = 1.76 (1.51, 2.07; p<0.001). However, households with CHE at 25% and 40% experienced relatively lower odds (compared to 10% of household income) of food insecurity, OR (95% CI; p) = 1.68 (1.43, 1.98; p<0.001) and OR (95% CI; p) = 1.69 (1.44, 1.99; p<0.001), respectively even though the odds in both cases were higher than households that did not experience CHE (Table 5).

**Table 5:** Bivariate relationships between CHE at different levels and household food insecurity.

| Characteristic | N | OR<sup>l</sup> | 95% CI<sup>l</sup> | p-value |
| --- | --- | --- | --- | --- |
| CHE at 10% | 3,424 |  |  |  |
| No |  | — | — |  |
| Yes |  | 1.76 | 1.51, 2.07 | <0.001 |
| CHE at 25% | 3,424 |  |  |  |
| No |  | — | — |  |
| Yes |  | 1.68 | 1.43, 1.98 | <0.001 |
| CHE at 40% | 3,424 |  |  |  |
| No |  | — | — |  |
| Yes |  | 1.69 | 1.44, 1.99 | <0.001 |

<sup>l</sup>OR = Odds Ratio, CI = Confidence Interval

### Predictors of CHE at various levels of income

In households where the household head had a high school education compared to households with the head not having a high school education, the odds of experiencing CHE was 1.16 (95% CI 0.99, 1.37; p=0.074) at 10%, 1.08 (95% CI 0.91, 1.27; p=0.4) at 25%, and 1.05 (95% CI 0.89, 1.25; p=0.6) at 40%, with none of the odds being statistically significant. In households where the head had post-high school education, the odds of CHE across all thresholds were lower compared to households where the head did not have post-secondary education: 0.61 (95% CI 0.46, 0.81; p<0.001) at 10%, 0.60 (95% CI 0.45, 0.79; p<0.001) at 25%, and 0.58 (95% CI 0.43, 0.77; p<0.001) at 40%. Households with food insecurity were significantly more likely to experience CHE, 1.58 (95% CI 1.34, 1.86; p<0.001) at the 10% of household income, but decreased to 1.49 (95% CI 1.26, 1.76; p<0.001) at 25%, and 1.48 (95% CI 1.25, 1.76; p<0.001) at 40%. Additionally, households with members aged over 60 years had increased odds of 1.45 (95% CI 1.22, 1.71; p<0.001) at the 10% threshold, decreased to 1.48 (95% CI 1.25, 1.76; p<0.001), at 25% and 1.44 (95% CI 1.21, 1.71; p<0.001) at 40% thresholds. Similarly, households with members under 18 years old had higher odds of CHE, with an OR of 1.32 (95% CI 1.09, 1.61; p=0.004) at 10% threshold but increased similarly in odds to 1.32 (95% CI 1.09, 1.61; p=0.006) at 25% and 1.33 (95% CI 1.09, 1.63; p=0.005) at 40%. Households in rural areas were more likely to experience CHE compared to those in urban areas, 1.47 (95% CI 1.25, 1.72; p<0.001), 1.32 (95% CI 1.09, 1.61; p=0.006), and 1.33 (95% CI 1.09, 1.63; p=0.005) at 10%, 25%, and 40%, respectively, relative to household income. Households in rural areas were more likely, 1.47 (95% CI 1.25, 1.72; p<0.001) at 10%, 1.53 (95% CI 1.30, 1.80; p<0.001) at 25%, and 1.60 (95% CI 1.35, 1.89; p<0.001) at 40% of household income, to experience CHE, all statistically significant (Table 6).

**Table 6:**
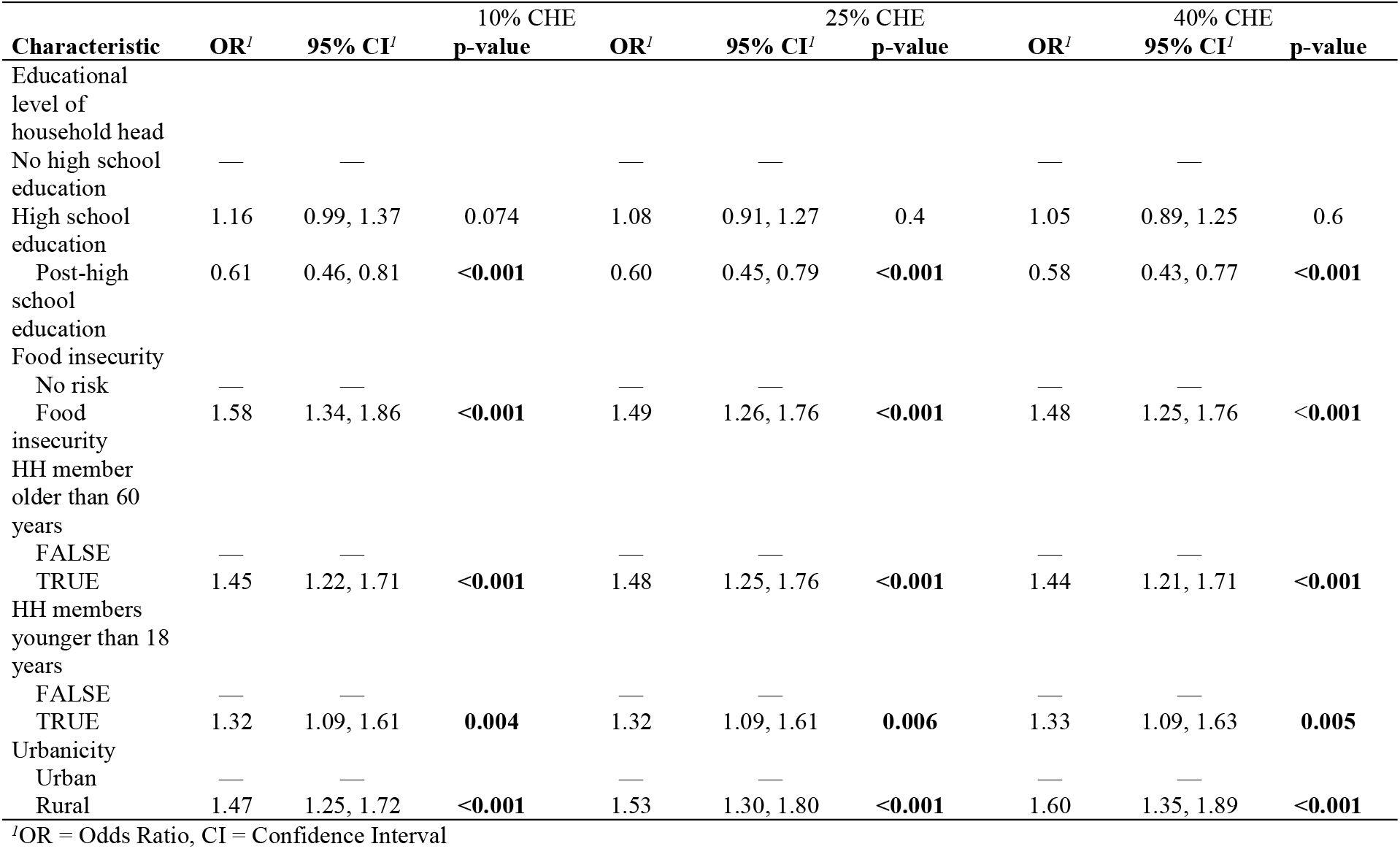
Predictors of household catastrophic health expenditure at 10% of total household income.

## Discussions

From our analysis, the mean household OOP for healthcare was 5.41 (49.66) (About half a dollar in 2023); adding travel costs and other direct charges, the mean of the total household health spending was 158.84 (316.44) and the equivalent of USD 14.18. Our analysis shows a 250% increase in OOP compared to a previous study (24), even accounting for time and inflation. The biggest proportion of the expenses was buying medicines and medical supplies Table 3. In all, 985 (16.4%) households had experienced CHE relative to 10% of household income. Our findings are consistent with the SSA estimate between 12.9% and 20.4% (6). Our CHE estimate, however, contrasts with the findings of Xu (2003). However, it must be noted that he relied on the Ghana Living Standards Survey in 1998 and, as such, can be deemed less reliable in the present context. Our findings also contrast with studies in Ethiopia. Still, it must be noted that social health insurance in Ghana started a decade earlier, and thus, the Ghana NHIS may be more mature than in Ethiopia. (45). Estimates for CHE at 25% and 40% of household income are similar, 929 (15.5%) and 903 (15.1%), respectively. The association between experiencing food insecurity and CHE is high across all levels (Table 5) and consistent with results in other studies. (17,42,46).

Our findings on the predictors of CHE are consistent once again with studies from other parts of the world. The educational level of the household head was found to protect against CHE if they had a post-high school education. We speculate that with higher education, household heads may be trying home over-the-counter medications, thereby not using the full range of health services. People with higher education may also be more likely to comply with health advice, averting illness and hospitalisation. Consistent with the available literature, having a household member older than 60 years or younger than 18 increased the likelihood of CHE by a factor of 1.45 and 1.32, respectively. Such household members need and consume more health services and are, therefore, more likely to push poorer households into catastrophic health expenditures. The problem may be compounded as these household members are not expected to be economically active, are in school, or are under apprenticeship. We found CHE to vary with urbanicity, which is consistent with other studies. (19).

## Study limitations

The study relied on secondary data collected by the Ghana Statistical Service (GSS). While GSS relied on trained enumerators (data collection assistants), the data is limited by the study respondents’ ability to sufficiently recall events that would have happened two weeks prior to the data collection.

## Conclusion

This paper shows a relatively high prevalence of Catastrophic Health Expenditure (CHE) in the Eastern region of Ghana. The prevalence of CHE is significantly associated with food insecurity at the 10%, 25%, and 40% thresholds of household income. Other CHE predictors are the household head’s educational level, the presence of a minor or older household member, and urbanicity, which impose an additional risk of a CHE. The national health insurance scheme is insufficient in providing financial risk protection, and the government must revisit its implementation. The government should consider enrolling more poor households into social programs like the Livelihood Empowerment Against Poverty (LEAP) to cushion them against CHE and food insecurity.

